# Transmission of SARS-CoV-2 among children and staff in German daycare centers: results from the COALA study

**DOI:** 10.1101/2021.12.21.21268157

**Authors:** Julika Loss, Juliane Wurm, Gianni Varnaccia, Anja Schienkiewitz, Helena Iwanowski, Anne-Kathrin Mareike Loer, Jennifer Allen, Barbara Wess, Angelika Schaffrath Rosario, Stefan Damerow, Tim Kuttig, Hanna Perlitz, Anselm Hornbacher, Bianca Finkel, Carolin Krause, Jan Wormsbächer, Anna Sandoni, Ulrike Kubisch, Kiara Eggers, Andreas Nitsche, Aleksandar Radonic, Kathrin Trappe, Oliver Drechsel, Kathleen Klaper, Andrea Franke, Antje Hüther, Udo Buchholz, Walter Haas, Lothar H. Wieler, Susanne Jordan

## Abstract

**Background:** Whereas the majority of children under 6 years of age attend daycare centers in Germany, evidence on the role of daycare centers in the transmission of SARS-CoV-2 is scarce.

**Aims:** This study aims to investigate the transmission risk in daycare centers among children and staff and the spread of infections to associated households.

**Methods:** 30 daycare groups with at least one recent laboratory-confirmed SARS-CoV-2 case (child or staff) were enrolled in the study (10/2020-06/2021). Close contacts within the daycare group and households were examined over a 12-day period (repeated SARS-CoV-2 PCR tests, genetic sequencing of viruses, documentation of symptoms). Households, local health authorities and daycare staff were interviewed to gain comprehensive information on each outbreak. We determined primary cases for all daycare groups.

**Results:** The number of secondary cases varied considerably between daycare groups. The pooled secondary attack rate (SAR) across all 30 daycare centers was 9.6%. The SAR tended to be higher in daycare centers in which the Alpha variant of the virus was detected (15.9% vs. 5.1% with evidence of wild type). The SAR in households was 53.3%. Exposed children were less likely to get infected with SARS-CoV-2 in daycare centers, compared to adults (7.7% vs. 15.5%).

**Conclusion:** Containment measures in daycare programs are critical and become increasingly important with highly transmissible new variants to reduce SARS-CoV-2 transmission, especially to avoid spread to associated households. Virus variants may modify transmission dynamics in daycare programs.

## Background

Since the beginning of the COVID-19 pandemic in March 2020, the role of daycare children in the spread of SARS-CoV-2 has been discussed controversially. In daycare centers children have close contact with each other and staff, possibly facilitating the transmission of SARS-CoV-2. Effective containment measures, such as physical distancing and wearing masks, are difficult to implement in early childhood. As of November 2021, COVID-19 vaccines have not been approved in Germany for application in toddlers and preschoolers yet. Given the high proportion of young children attending daycare (in Germany, 35% of 0- to 2-year-olds and 93% of 3- to 6-year-olds, [1]), understanding SARS-CoV-2 transmission within daycare centers is critical to inform adequate mitigation policies.

Only few studies have explored transmission dynamics in daycare centers or among children of kindergarten age. An early publication from Australia found that children were the primary case in only three of ten daycare centers with SARS-CoV-2 cases; in none of these daycare centers a secondary case was detected among close contacts [2]. According to a review by Spielberger et al., children with SARS-CoV-2 infected an average of 13.4% of their contacts [3], whereas more recent meta-analyses on SARS-CoV-2 report transmission rates of 4% and (approx.) 5% if children were registered as index case [4, 5]. These results cannot be transferred to daycare centers, since the analyses referred to broader age groups (0-9 years and older), and the data primarily stem from household studies, not from daycare programs. A meta-analysis of contact tracing studies concluded that children are less susceptible than adults to SARS-CoV-2, but with large heterogeneity between studies [6]. The data basis on transmission risk of children in daycare centers is therefore still unsatisfactory. In addition, COVID-19 in childhood often goes along with no or only mild symptoms [7], which may obscure the role of children in transmission dynamics, as they may not be documented as index case in an outbreak. Transmission dynamics may also be influenced by virus variants. An increased transmissibility was postulated with the advent of SARS-CoV-2 “variants of concern” (VOC), such as the Alpha variant [8], although its role in outbreaks in daycare centers remains unknown.

When SARS-CoV-2 began to spread in Germany in March 2020, several mitigation measures were implemented, such as physical distancing and hygiene measures. Daycare programs were temporarily suspended, or restricted to a limited number of children (who had special needs, or whose parents had critical jobs). Furthermore, different containment measures were implemented within daycare centers, i.e. hygiene plans, taking children’s temperature, or regular ventilation. The study presented in this paper was carried out during the “second wave” (10/20–02/21) and “third wave” (03/21–05/21) of the pandemic in Germany, during which numerous outbreaks occurred in daycare centers (between ca. 50-200 per week nationwide) [9]. Due to the timing of this study it was not possible to investigate the role of more recent variants of concern, like Delta, as those emerged later in Germany.

In order to gain a better understanding about the transmission risk of SARS-CoV-2 within daycare centers and corresponding households of infected study participants, we conducted an observational study in German daycare centers with at least one notified case of SARS-CoV-2 (“Corona outbreak-related examinations in daycare centers”, COALA). The aim of the study was to identify secondary infections among exposed children and adults in daycare centers and their corresponding households, the primary endpoint being the secondary attack rate (SAR) in both settings.

## Methods

### Study design

Design and methods of COALA are described in detail in a study protocol [10]. We chose a case-ascertained study design with longitudinal collection of data and specimen. We included daycare center groups in which one or more SARS-CoV-2 positive cases (child or staff) were detected, and parents and staff consented to participate. From October 2020 till June 2021, 30 daycare groups from 20 different communities all over Germany were included in the study. SARS-CoV-2 cases as well as their close contacts within the respective daycare group and household were examined over a 12-day period, including the collection of biological specimens and the documentation of symptoms. In order to reconstruct transmission chains and identify primary cases, clinical symptoms and possible exposition towards the virus were assessed retrospectively through questionnaires. The SARS-CoV2-antibody status was assessed in capillary blood samples to detect previous infections.

### Recruitment

Information about newly diagnosed SARS-CoV-2 cases in daycare centers was gathered through collaboration with several local health authorities, or contacts with (umbrella) associations that run daycare facilities. The recruitment was restricted to daycare centers in which the participants could be seen and tested by the study personnel within 4-6 days after the PCR test of the index case; it also followed a purposive sampling strategy, to allow for a roughly equal distribution of children and adults among index cases. Index cases and their close contacts (within the daycare group) were asked to participate, both children and adults. Household members of index cases and infected close contacts were included as well. Written informed consent was obtained from each participant. Participants received a monetary incentive, in order to increase adherence to the complex and time-consuming self-sampling schedule.

### Procedure of collecting data and biological samples

As study participants were isolated or quarantined, they were visited at home by trained study personnel within four to six days after the PCR test of the index case. This timeframe was chosen based on the mean incubation time of SARS-CoV-2 [11], and in order to ensure the detection of secondary cases. We took combined mouth and nose swabs, saliva samples and capillary blood samples on dried blood spot cards from each participant. Participants were also instructed to take combined mouth and nose swabs and saliva samples from themselves and/or their children in a cycle of three days (over a total period of 12 days) as well as to return their samples independently to the laboratory via mail (Figure 1). Mouth and nose swabs and saliva samples were chosen instead of nasopharyngeal swabs because they are more suitable for self-testing and testing of children, respectively, while presenting convincing sensitivity and specificity [12-14].

**Figure 1:**
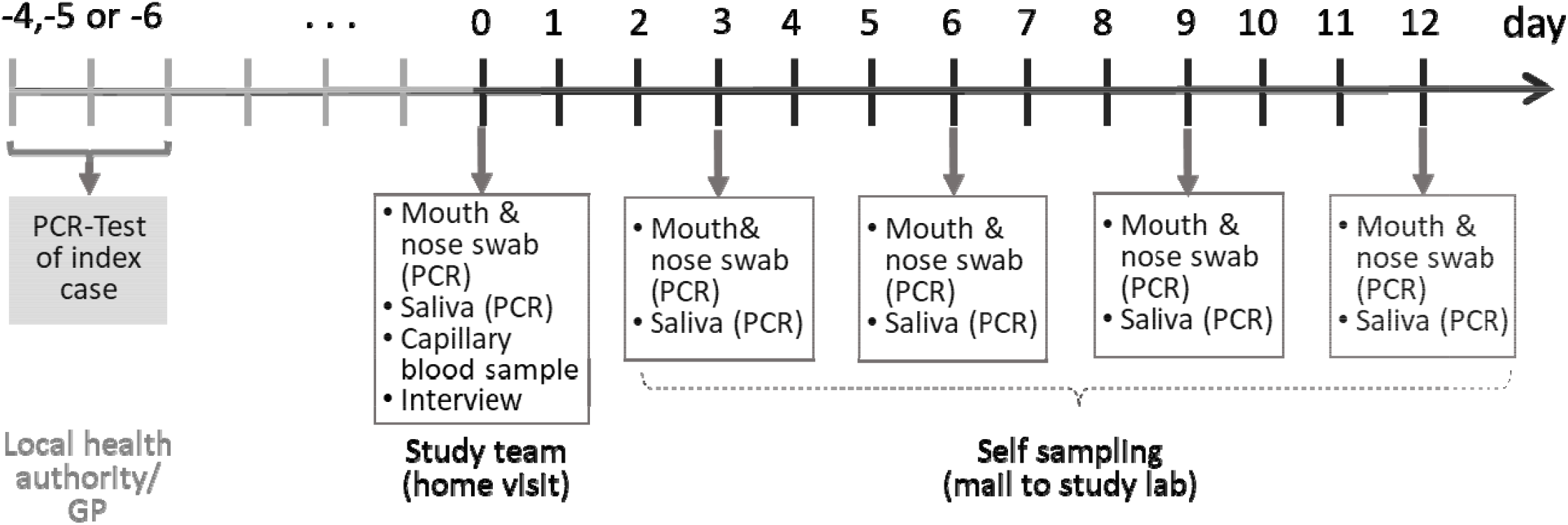
Time schedule for bio samples taken from the participants of the COALA study (SARS-CoV-2 index cases, secondary cases and close contacts of SARS-CoV2 cases in the respective daycare center group and households). Participants were enrolled 4-6 days after the index case got tested. GP = general practitioner

### Laboratory testing

Combined mouth and nose swabs and saliva samples were tested for viral RNA of SARS-CoV-2 by real-time reverse-transcriptase-polymerase chain reaction (rRT-PCR). Positive samples were also screened for the Alpha variant by PCR, and the genome was sequenced to detect virus lineages [15]. Sequencing was done on Illumina ISeq (2x 150bp; CleanPlex amplification) or ONT MinION devices (ARTIC v3 amplification [16]). Virus sequences were determined using covpipe (Illumina data, [17]) or poreCov (MinION data, [18]). Capillary blood samples were tested for IgG antibodies against the S1 domain of SARS-CoV-2 spike protein with a semiquantitative Enzyme-Linked Immunosorbent Assay (ELISA from Euroimmun, Germany).

### Questionnaires

Standardized telephone interviews were conducted with each participating household, to gain information on each household member regarding a previous SARS-CoV-2 infection, SARS-CoV-2 exposition, history of COVID-19 symptoms, and the children’s attendance times at the daycare center. Further information on the outbreak was gained through interviews with the respective local health authorities and daycare director.

### Definitions

All participants from the included daycare groups (SARS-CoV-2 cases and close contacts) constituted the daycare center cohort. The household cohort comprised the secondary cases of the daycare group (children or staff members) and their respective household members. The index case was defined as the first case of SARS-CoV-2 to be notified and reported to the local health authority, whereas the primary case was considered to be the origin of the infectious event in the daycare group or household. Primary case and index case were not necessarily identical with each other. A secondary case was defined as a close contact of the primary case who tested positive in a SARS-CoV-2 PCR test up to 9 days after the household visit. Close contacts were defined as individuals exposed to the confirmed SARS-CoV-2 cases within the daycare group or household, respectively. Secondary cases infected in the daycare center were considered to be primary cases in their respective households. The secondary attack rate (SAR) is the proportion of infected contacts out of the total number of (susceptible) contacts within a setting.

### Determination of primary cases

Based on the results of the laboratory testing and of the standardized interviews, we reconstructed infection pathways in each household and daycare group, and thereby determined the probable primary cases. The probable date of infection with SARS-CoV-2 was estimated for every participant with SARS-CoV-2 following the proceedings suggested by Layan et al. [19], considering (a) information on symptom onset and (b) date of first positive PCR test. Furthermore, we considered information on exposure to SARS-CoV-2 and IgG-antibody status against SARS-CoV-2.

Probable primary cases were determined for all investigated daycare groups. In four daycare groups, there was convincing evidence that an individual other than the index case had been the first person to be infected within the group, mainly due to earlier onset of symptoms, or exposition to an earlier laboratory-confirmed COVID-19 infection in the household, or both. In two of those four daycare groups, a child was determined as the probable primary case instead of a reported adult index case. There was also a slight change in the number of close contacts (plus two), as in two daycare groups where two children (siblings)/staff had been registered as simultaneous index cases, another child was determined as the probable primary case.

### Statistical analyses

Descriptive statistics were performed to describe secondary cases and secondary attack rates. Confidence intervals were calculated on the logit scale using robust standard errors, accounting for the clustering within daycare groups or households, using the Stata survey command. Odds ratios (OR) and *p*-values were on a GEE (generalized estimating equations) logistic regression model [20] with exchangeable working correlation to account for clustering. Since the number of clusters was small, a Mancl-de Rouen correction was applied [20-23]. All analyses were performed using Stata 17.0.

### Sample characteristics

Between October 2020 and June 2021, 85 daycare centers with an acute SARS-CoV-2 case were reported to the study team. Out of 85 these, 30 daycare groups were included in the COALA study. Reasons for not including a daycare center were a lack of research capacity (i.e. in case of a high number of simultaneously reported outbreaks), insufficient response among potential participants, or prioritization of daycare centers with a child as index case. In 17 of the 30 daycare groups, the index case was reported to be a child (or two children, who were siblings and tested positive on the same day, n=3), in 13 daycare groups it was a staff member.

Overall, 282 daycare children (1-6 years), 91 staff members (19-68 years) and 45 household members (1-69 years) of secondary cases in the daycare groups were included in this analysis (Tab. 1). In several daycare groups, not all members of the included daycare groups consented to participate in the study; the response rate among index cases was 74%, among close contacts 60% (children) and 57% (staff), respectively.

IgG antibodies against SARS-CoV2 were detected in 22 cases (children: n=8, staff: n=12; adults in households n=2); 6 had a simultaneous positive PCR-test and no history of prior infection, and were determined to be in a state of fresh seroconversion (3 of them were primary cases in the daycare group).

## Results

### Secondary transmissions in daycare groups

We detected 33 secondary SARS-CoV-2-cases during the course of the study through PCR testing (Tab. 2 and 3); 3/33 were not detected before the self-sampling phase. 6 additional secondary cases were close relatives (e.g. siblings) of the primary cases who attended the same daycare group; they were excluded from the analysis, as it could not be determined whether they got infected during daycare or at home. The number of secondary transmissions varied considerably among daycare groups. In the majority of the daycare groups, no secondary cases (22/30) or only 1-2 secondary cases (3/30) were detected among the participating close contacts. The maximum number of secondary attacks observed in one daycare group was 11.

### Secondary attack rate in daycare groups

When the results of all daycare groups were pooled, the mean SAR in the daycare cohort was estimated at 9.6% (95% CI: 4.0–21.3%) (Tab. 4). When close contacts with existing SARS-CoV-2 antibodies (n=17) were excluded from the calculation due to an assumed lower susceptibility, the attack rate remained similar (9.5%, 95% CI: 4.1–20.6%). The transmission risk from pediatric primary cases did not differ significantly from that from adult primary cases (11.2% vs. 7.0%, *p*=0.706). When excluding a daycare group with extraordinarily high transmission of 11 secondary cases (no. 24), the estimation of the SAR from children decreased from 11.2 to 6.5% (95% CI: 1.7-21.5%).

Most studies do not determine primary cases, but calculate the SAR based on the reported index cases. To investigate whether the determination of primary cases made a difference to the SAR, we also estimated it based on index cases. In this case, the result for the transmission risk from infected children to close contacts would have been different: 4.2% (95% CI: 0.7–20.6%) instead of 11.2%, versus 17.3% (95% CI: 6.2– 39.9%) instead of 7.0% from adult index cases (staff) (*p*=0.095). This could be explained by the fact that those two daycare groups where the determined primary case was a child, whereas the index case reported to the health authority had been an adult, were no. 11 and no. 24. These were two outbreaks with a relatively high number of secondary cases (n=4 and n=11).

In daycare groups with a confirmed infestation of the Alpha variant (n=15), the estimated SAR was higher than the SAR for groups with confirmed infections of the wild-type or another non-VOC (5.1% vs. 15.9%), but not statistically significant. Regarding susceptibility towards the virus, children in the daycare groups were less likely to have contracted SARS-CoV-2 than staff. 7.7% of all participating children who were exposed to SARS-CoV-2 within their daycare group tested positive for SARS-CoV-2, compared to 15.5% of all adult close contacts (*p*=0.005).

### Secondary attack rate in households

Those children (n=20) and staff members (n=13) who got infected with SARS-CoV-2 during daycare lived in 24 households with 45 close contacts (34 adults, mostly parents, and 11 children, mostly siblings). From those, two single-person households of infected staff members were consecutively excluded from further analysis. In six of the remaining 22 households, two or more infected daycare children or staff members lived together in one household. Overall, secondary infections among further household members were observed in 12 of the 22 households (54.5%). In total, 24 of all 45 close contacts in households were diagnosed as secondary SARS-CoV-2 cases, resulting in a mean household SAR of 53.3% (35.4–70.4%), the SAR being significantly higher in households than in the daycare setting (*p*=0.000).

## Discussion

### Summary of main findings

Across the 30 studied daycare centers with a SARS-CoV-2 case, the transmission risk was very heterogeneous. In most daycare groups, there were no transmissions among participants, whereas there were few daycare groups in which we detected numerous secondary cases. When pooling the daycare groups, we found a secondary attack rate of 9.6%, which was significantly lower than the SAR in the associated households (53.3%). A higher proportion of infections among close contacts was found in daycare groups with the Alpha variant, as compared to wild-type or non-VOC (15.9% vs. 5.1%). There was no significant difference in the secondary attack rate when children were the primary case in comparison to when adults were the primary case, but exposed adults had a significantly higher risk of contracting SARS-CoV-2 than children. In case of an infection in the daycare group, the virus was introduced to 54.5% of corresponding households.

### Comparison with other studies

A comparable study was conducted in one federal state of Germany, using data from routine contact tracing. It included 99 index cases from daycare centers and 4392 contacts in daycare [24], with a resulting SAR of 2.5%. As the study was performed earlier (from August to December 2020), it could not include cases with the Alpha variant, which may account for the lower SAR in this study as compared to our analysis (9.6%); the SAR of non-VOC outbreaks in our study was 5.1%. The follow-up period of 12 days after enrollment and regular testing could identify additional cases in COALA, who otherwise might have not been detected in routine contact tracing due to mild or absent symptoms, which could also explain the higher SAR found in our study.

### Implications for policy, practice, and research

Our results thus confirm that SARS-CoV-2 transmissions occur in daycare center groups, and that both children and staff members can play a role in infectious events. The predominance of the Alpha variant resulted in a higher transmission risk in daycare (although statistically not significant). This shows that the role of children, or daycare centers, may change considerably over the course of the pandemic, depending on the predominant genetic lineage. This confirms the need for continuous monitoring of outbreaks and transmission patterns in this setting. Still, given the fact that physical distancing and wearing of masks is hardly possible among toddlers and preschoolers within a daycare center group, it is striking that the SAR in this setting is still significantly lower than in households (9.6% vs. 53.3%). Even if only a relatively small percentage of close contacts gets infected with SARS-CoV-2 in the daycare group, those who were infected were quite probable to spread the infection to their families and household members. Daycare centers may have the potential to serve as bridges over which the virus can be spread to other households. Daycare providers can therefore help slow the spread of COVID-19 by implementing mitigation measures in their programs. One of the most effective approaches is keeping the groups within daycare centers small and consistent, and minimizing contact between these groups [25]. The role of different COVID-19 testing strategies is still to be evaluated.

When analyzing the SAR within a specific setting, or from a specific population (e.g. children), it is worth determining the primary case where possible, even when analyzing pooled samples. Otherwise, the role of children in outbreaks may be underestimated.

### Strengths and limitations

The study is subject to some limitations. First, the sample size is relatively small, which is why the results show a relatively high statistical uncertainty. Furthermore, sampling of daycare centers was not random and might not be representative. In addition, with the study being voluntary, the participation among close contacts in daycare groups and households was mostly not complete. We cannot rule out a selection bias, for example severely ill persons may have declined more frequently than others the invitation to participate. When determining the probable primary case, we were sometimes also restricted by the fact that not all daycare group members participated, so we could not rule out that maybe the real primary case would be found outside of our sample. Including additional data from local health authorities and daycare directors was meant to compensate for this drawback.

A strength of our study is that we were largely successful in determining primary cases. The reconstruction of the transmission chain in each participating daycare group yielded probable primary cases which differed from the registered index cases in four of the daycare groups, twice with a change from staff member to child, thereby producing a different SAR when stratified for child vs. adult.

The prospective design was a strength of the study, as it enabled us to detect additional secondary cases which might have not been noticed if the investigation had taken place only briefly after the diagnosis of the index case. A further strength is the richness of data, including measurement of antibodies and close monitoring of symptoms, which is unusual in the non-clinical setting, and helped us track transmission dynamics and determine primary cases. By determining probable primary cases, we decreased the risk that children with mild or no symptoms may be overlooked as an infectious source in the daycare centers. Genomic sequencing of SARS-CoV-2 was a further advantage of the study, as the existence of a VOC (Alpha), which was found in 50% of included daycare group outbreaks, could partly explain the heterogeneity in the number of secondary cases between daycare groups. There was a lack of (sufficient) material to perform genomic sequencing in five daycare groups, however, which was favored by the fact that no or few participants were tested positive, so we cannot rule out that the SAR for daycare groups with the Alpha variant was overestimated.

A strength of the analysis is that the clustering of contacts within daycare groups and households, respectively, was taken into account when calculating confidence intervals and *p*-values, and that the effect of heterogeneity in SAR estimates across daycare groups on the results was explored.

## Conclusion

The COALA study adds to the global literature on SARS-CoV-2 by providing evidence on virus transmission in the daycare center setting, which has been understudied so far. Studies may risk to underestimate the secondary attack rate of children if they refer to reported index cases instead of determining probable primary cases, or implement only short follow-up periods. The study also shows that defining the SAR of SARS-CoV-2 cases in children is a moving target, as it may change over time with novel genetic variants of the virus and with varying range of mitigation measures that are put into place in daycare centers. Therefore, continuous monitoring of outbreaks and transmission patterns in daycare centers can contribute to developing, implementing and adapting adequate prevention and mitigation measures. Reducing transmission in daycare centers is critical, as children and staff members who got infected in the daycare group have a high risk of spreading the virus to their households.

## Data Availability

The authors confirm that some access restrictions apply to the data underlying the findings. The data set cannot be made publicly available because informed consent from study participants did not cover public deposition of data. However, the minimal data set underlying the findings is archived in the Health Monitoring Research Data Centre at the Robert Koch Institute (RKI) and can be accessed by researchers on reasonable request. On-site access to the data set is possible at the Secure Data Center of the RKIs Health Monitoring Research Data Centre. Requests should be submitted to the Health Monitoring Research Data Centre, Robert Koch Institute, Berlin, Germany.

## Ethical statement

The Ethics Committee of the Berlin Medical Association has reviewed the COALA study and approved the implementation of the study (Eth-39/20). The participation in the study is voluntary and all participants are informed about the objectives and contents of the study as well as data protection and give their written informed consent; children aged from 14 to 17 years give their own written consent in addition. Every participant is assigned to a sequential study number (ANR) in order to ensure pseudonymization of the study documents and material.

COALA is registered in the German Clinical Trail Register (DRKS-ID=DRKS00023501).

## Funding

Corona outbreak-related examinations in daycare centers – COALA is conducted by the Robert Koch Institute (RKI) and is funded by the Federal Ministry of Health under grant number ZMVI1-2520COR404 for the period June 2020 to December 2021. COALA is one out of four modules of the “Corona daycare center study: Research on the organizational, hygienic and educational challenges of emergency daycare in daycare centers as well as on acute respiratory diseases during the implementation of measures to contain SARS-CoV-2”, which is being conducted by the German Youth Institute (DJI) together with the RKI. The Federal Ministry of Health was not involved in the design of the study or collection of data.

## Contributions

JL, ASch, UB, WH, HP, AHo, LHW and SJ designed the study. JWu, HI, ASa, UK, JWo, BF, CK and KE were responsible for participant recruitment, data and specimen collection. ASa, UK, AN performed laboratory testing and analysed lab results, AR, KT, OD and KK were responsible for genome sequencing and genome reconstruction, A-KL, JA, BW, AF, AHü, TK designed the databases and managed and linked the data. WH, UB, JL, GV, Asch, AHo and SJ conceptualized the analysis. GV, JWu, ASch, ASR, SD carried out the analysis and were responsible for the accuracy of the data analysis.

JL, JWu, GV, ASch, HI, BF, CK and SJ wrote the manuscript, which was then reviewed and approved by all other authors.

## Informed consent

Written informed consent to participate in this study was provided by the participants or their legal guardian, respectively.

## Data access

Genome sequences determined within this study are published in ENA/GISAID “StudyID”.

## Acknowledgements

We would like to thank all our colleagues at the Robert Koch Institute in the Department Epidemiology and Health Monitoring, the Department Infectious Disease Epidemiology, the Centre for Biological Threats and Special Pathogens, and the sequencing unit at MF2 for their support. Special thanks to the colleagues of SurvAd, the Epidemiological Central Laboratory, and the COALA study team at the Robert Koch Institute. We would like to thank all participants for their support of the study. For their advice in the development of the study design we would like to thank all federal state health authorities, local health authorities, experts on daycare centers, and to colleagues at the German Youth Institute (DJI) who supported us designing the interviews.

---

**Table 1:** Characteristics of the study population in daycare groups and households with at least one acute laboratory-confirmed SARS-CoV-2 case, COALA study, Germany, 10/2020–06/2021

|  | Children and adults (staff) in the daycare groups with at least one SARS-CoV-2 case |  |  | Household members of the secondary cases in the daycare groups |  |  |
| --- | --- | --- | --- | --- | --- | --- |
|  | Children | Adults | Total | Children | Adults | Total |
| **Sex** |  |  |  |  |  |  |
| Female | 128 | 82 | 210 | 8 | 17 | 25 |
| Male | 154 | 9 | 163 | 3 | 17 | 20 |
| **Age** |  |  |  |  |  |  |
| 1-6 years | 282 | - | 282 | 2 | - | 2 |
| 7-17 years | - | - | - | 9 | - | 9 |
| 18+ years | - | 91 | 91 | - | 34 | 34 |
| **Role** |  |  |  |  |  |  |
| Primary case<sup>a</sup> | 16 | 7 | 23 | - | - | - |
| Close contact<sup>b</sup> | 266 | 84 | 350 | 11 | 34 | 45 |
| **Total** | 282 | 91 | 373 | 11 | 34 | 45 |

<sup>a</sup>In 7 daycare groups, the primary cases were tested for SARS-CoV-2 and officially confirmed by the local health authorities, but did not participate in our study. Nevertheless, these daycare groups were included, as a substantial number of close contacts consented to participate, which was essential for calculating the secondary attack rate.
<sup>b</sup>Including 7 siblings of primary cases in the daycare group that are excluded in further analysis because it cannot be determined whether they got infected in the daycare group or their household.

**Table 2:** Characteristics of the daycare groups with at least one acute laboratory-confirmed SARS-CoV-2 case, COALA study, Germany, 10/2020–06/2021

| Daycare group | Month of outbreak | Type of primary case | Number of participating close contacts<sup>a</sup> | Number of secondary transmissions | Secondary attack rate | Number of close contacts with existing antibodies at day 0 | Virus variant |
| --- | --- | --- | --- | --- | --- | --- | --- |
| 1 | 10/20 | Child | 13 | 0 | 0.0 | 0 | Wildtype /non-VOC |
| 2 | 10/20 | Adult | 9 | 1 | 11.1 | 0 | Wildtype /non-VOC |
| 3 | 11/20 | Adult | 18 | 0 | 0.0 | 0 | Wildtype /non-VOC |
| 4 | 11/20 | Child | 14 | 0 | 0.0 | 0 | Wildtype/non-VOC |
| 5 | 12/20 | Child | 12 | 0 | 0.0 | 0 | Wildtype/non-VOC |
| 6 | 01/21 | Adult | 23 | 0 | 0.0 | 1 | Unknown |
| 7 | 01/21 | Child | 13 | 1 | 7.7 | 0 | Wildtype/non-VOC |
| 8 | 01/21 | Child | 7 | 0 | 0.0 | 0 | Unknown |
| 9 | 01/21 | Adult | 8 | 0 | 0.0 | 1 | Wildtype/non-VOC |
| 10 | 02/21 | Adult<sup>b</sup> | 12 | 3 | 25.0 | 0 | Wildtype/non-VOC |
| 11 | 02/21 | Child<sup>bc</sup> | 11 | 4 | 36.4 | 0 | VOC-Alpha |
| 12 | 02/21 | Child | 9 | 0 | 0.0 | 0 | VOC-Alpha |
| 13 | 02/21 | Adult | 10 | 0 | 0.0 | 0 | VOC-Alpha |
| 14 | 02/21 | Child | 5 | 0 | 0.0 | 1 | Wildtype/non-VOC |
| 15 | 02/21 | Child | 15 | 0 | 0.0 | 0 | VOC-Alpha |
| 16 | 02/21 | Child | 18 | 0 | 0.0 | 0 | VOC-Alpha |
| 17 | 03/21 | Child | 7 | 0 | 0.0 | 0 | VOC-Alpha |
| 18 | 03/21 | Adult | 7 | 0 | 0.0 | 0 | Unknown |
| 19 | 03/21 | Child | 16 | 0 | 0.0 | 1 | VOC-Alpha |
| 20 | 03/21 | Adult | 13 | 1 | 7.7 | 0 | Wildtype/non-VOC |
| 21 | 03/21 | Child | 9 | 0 | 0.0 | 0 | Unknown |
| 22 | 03/21 | Child | 8 | 0 | 0.0 | 1 | VOC-Alpha |
| 23 | 03/21 | Adult | 14 | 4 | 28.6 | 0 | VOC-Alpha |
| 24 | 03/21 | Child<sup>bc</sup> | 14 | 11 | 78.6 | 1<sup>d</sup> | VOC-Alpha |
| 25 | 04/21 | Child | 7 | 0 | 0.0 | 2 | VOC-Alpha |
| 26 | 04/21 | Child<sup>b</sup> | 19 | 8 | 42.1 | 1<sup>d</sup> | VOC-Alpha |
| 27 | 05/21 | Adult | 7 | 0 | 0.0 | 1 | VOC-Alpha |
| 28 | 05/21 | Adult | 7 | 0 | 0.0 | 3 | VOC-Alpha |
| 29 | 06/21 | Child | 10 | 0 | 0.0 | 1 | Unknown |
| 30 | 06/21 | Child | 8 | 0 | 0.0 | 3 | VOC-Alpha |
| Total | - | - | 343 | 33 | 9.6<sup>e</sup> | 17 | - |

<sup>a</sup> 7 siblings of primary cases were excluded (6 of them were tested positive). <sup>b</sup> The primary case was not the index case. <sup>c</sup> The index case was an adult, the primary case a child. <sup>d</sup> The person was also tested positive on SARS-CoV-2. <sup>e</sup> Pooled secondary attack rate (33 of 343 participating close contacts were tested positive).

**Table 3:**
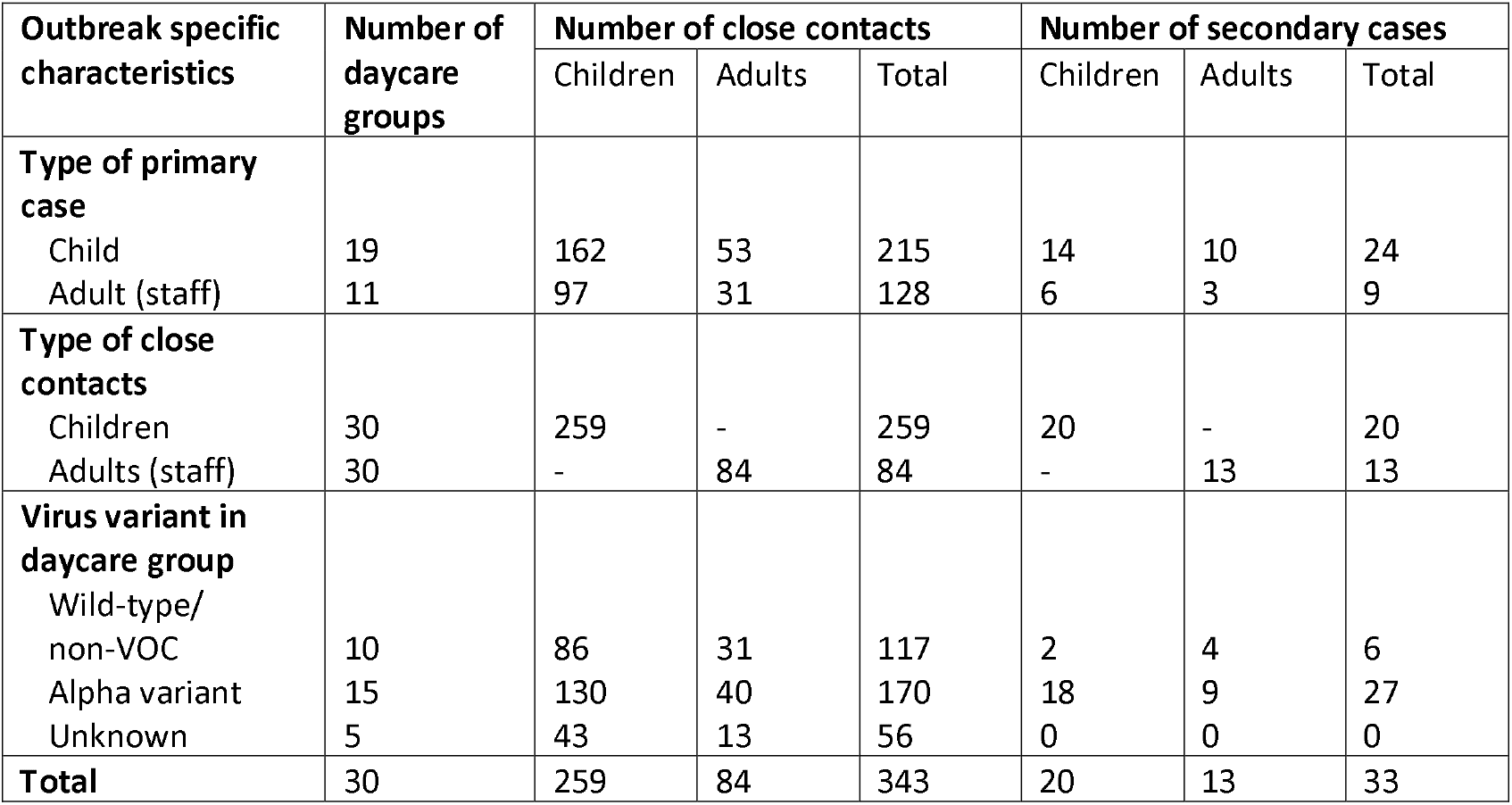
Close contacts and secondary cases in daycare groups with at least one acute laboratory-confirmed SARS-CoV-2 case by outbreak specific characteristics, COALA study, Germany, 10/2020–06/2021

**Table 4:** Secondary attack rates in daycare groups with at least one acute laboratory-confirmed SARS-CoV-2 case by outbreak specific characteristics, COALA study, Germany, 10/2020– 06/2021

| Outbreak specific characteristics | Secondary attack rate (95% CI)<sup>a</sup> | Unadjusted |  | Adjusted<sup>a</sup> |  |
| --- | --- | --- | --- | --- | --- |
|  |  | Odds ratio (95% CI) | p-value | Odds ratio (95% CI) | p-value |
| **Type of primary case** |  |  |  |  |  |
| Child | 11.2 (3.6–29.7) | 1.00 |  | 1.00 |  |
| Adult (staff) | 7.0 (2.5–18.5) | 0.73 (0.13–4.01) | 0.706 | 0.79 (0.13–4.82) | 0.794 |
| **Type of close contacts** |  |  |  |  |  |
| Children | 7.7 (2.7–19.9) | 1.00 |  | 1.00 |  |
| Adults (staff) | 15.5 (6.4–33.0) | **3.03** (1.45–6.32) | 0.005 | **3.11** (1.43–6.77) | 0.006 |
| **Virus variant in daycare group** |  |  |  |  |  |
| Wild type/<br>non-VOC | 5.1 (1.8–13.7) | 1.00 |  | 1.00 |  |
| Alpha variant | 15.9 (5.9–36.1) | 2.72 (0.52–14.4) | 0.225 | 1.97 (0.36–10.7) | 0.412 |
| **Total<sup>b</sup>** | 9.6 (4.0–21.3) | - | - | - | - |

<sup>a</sup> Adjusted for the variables in the first column. <sup>b</sup> The working correlation in the GEE model (intercept only) was estimated as 0.42 (excluding daycare group 24: 0.26). In the fully adjusted model, the working correlation was 0.36 (excluding daycare group 24: 0.22).

## Notes

### Competing Interest Statement

The authors have declared no competing interest.

### Clinical Protocols

https://www.frontiersin.org/articles/10.3389/fpubh.2021.773850/full

### Author Declarations

The Ethics Committee of the Berlin Medical Association has reviewed the COALA study and approved the implementation of the study (Eth-39/20). The participation in the study is voluntary and all participants are informed about the objectives and contents of the study as well as data protection and give their written informed consent; children aged from 14 to 17 years give their own written consent in addition. Every participant is assigned to a sequential study number (ANR) in order to ensure pseudonymization of the study documents and material. COALA is registered in the German Clinical Trail Register (DRKS-ID=DRKS00023501).

